# The BCG dilemma: linear versus non-linear correlation models over the time of the COVID-19 pandemic

**DOI:** 10.1101/2020.06.13.20129569

**Authors:** Guillermo Lopez-Campos, Christopher Hawthorne, Miguel A. Valvano

## Abstract

In this work we have analysed the use of two different correlation metrics and two different datasets, detecting differences in the extent and directionality of the correlation between COVID-19 infection and BCG immunization. Correlation between COVID-19 infection, death rates and BCG immunization is strongly affected by the correlation metrics and so will be their biological interpretation.

## Introduction

BCG immunization has been suggested to have a protective role for COVID-19 since, due to its immunomodulatory properties, it would appear to reduce the severity of other viral infections^1^. This was supported by several different analytical approaches and datasets analyzing the relationship between BCG vaccination rates around the world, COVID-19 cases and death rates^2-4^. Previous correlation analyses highlighted a negative correlation between BCG immunization and COVID-19, providing impetus for randomized clinical trials to evaluate if the BCG vaccine can reduce the severity of COVID-19^5^.

We have applied two different correlation methods (Spearman’s and Pearson’s) using the package psych in R, fortnightly COVID-19 infection and mortality rates between 15th January and 15th May 2020 (https://www.ecdc.europa.eu/en/publications-data/download-todays-data-geographic-distribution-covid-19-cases-worldwide), and two different BCG immunization datasets extracted from the WHO between 1980 and 2018 for one year old (1yo) children and undefined age immunization rates (https://www.who.int/data/maternal-newborn-child-adolescent/indicator-explorer-new/mca/baccille-calmette-gu%C3%A9rin-(bcg)-immunization-coverage-among-1-year-olds-(-) and https://www.who.int/immunization/monitoring_surveillance/data/en/).

The results show that in 1 year old children a negligible (or weakly positive) correlation between BCG immunization rates and COVID-19 death or case rates (15^th^ May 2020) using Spearman’s correlation. Whereas the opposite is observed using Pearson’s correlation with a statistically significant weak to moderate negative correlation. Spearman’s correlation allows the identification of monotonic trends compared to Pearson’s linear correlation. The latter as well tends to fail in skewed distributions and is more sensitive to extreme values such as those observed in some non-vaccinated countries. Similar opposite results were observed when we used the alternative BCG immunization rates (Figure 1A).

**Figure 1.**
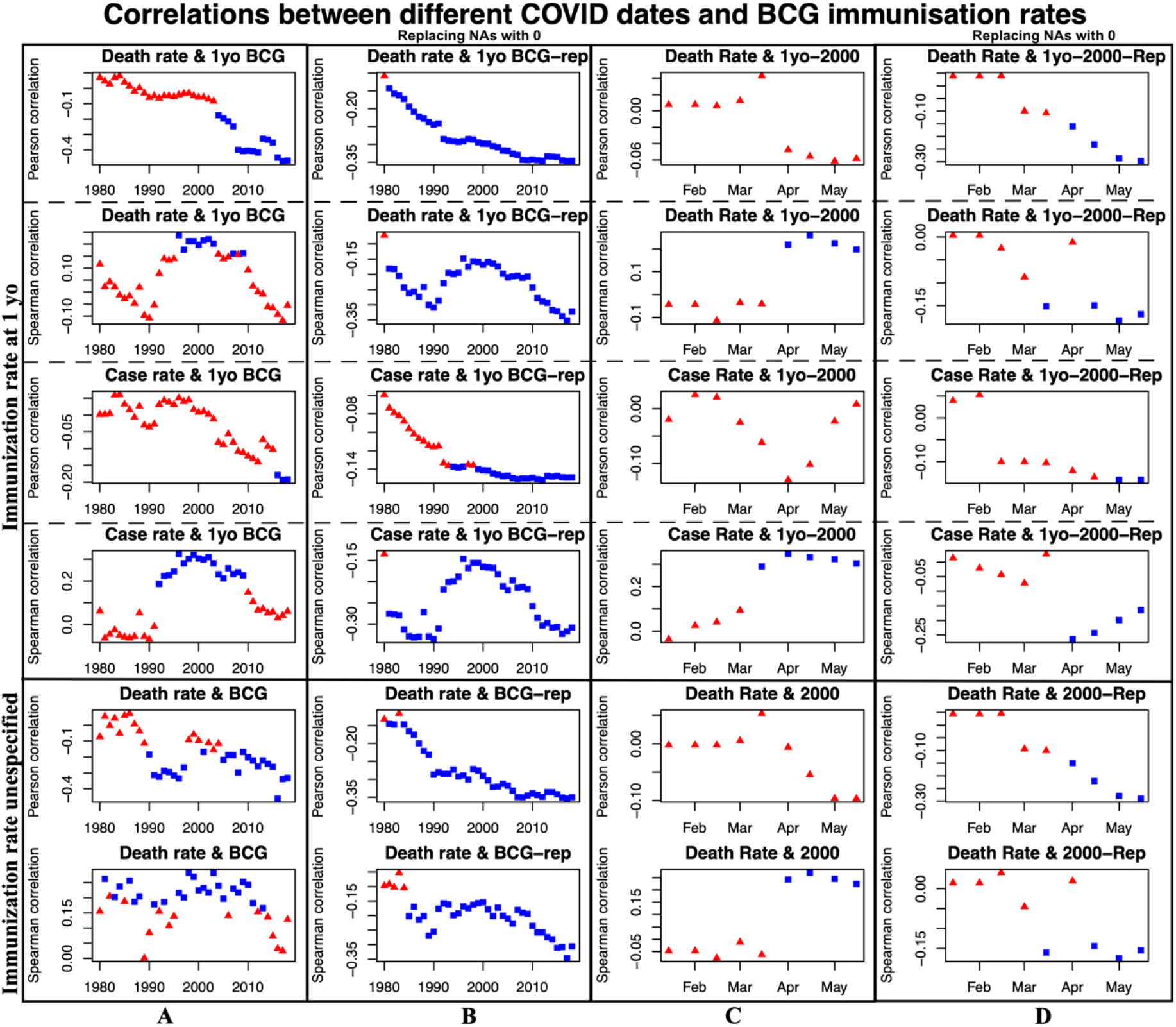
Correlation between BCG immunization rates and COVID-19 death and case rates. In blue statistically significant correlations (p-value <0.05)

We also analyzed fortnightly COVID-19 correlations during the pandemic and immunization rates in two different years (2000 and 2018). As before, Spearman’s correlation data show a weak positive correlation over time (Figure 1C). However, correlation increased until mid-April when many countries started implementing different protective measures, it proceeded to declined slightly and seems to have stabilized in May.

The datasets used for these analyses lack some countries that either had BCG immunization policy at later ages, did not have them or it was discontinued or non-reported (E.g. UK, USA and Australia). To address this aspect, we included an oversimplified replacement of missing BCG coverage data with a 0% coverage. This replacement introduces important changes in the results obtained, inverting the previously discussed results showing a negative correlation between death rates and COVID-19 for both correlation metrics (Figure 1B and 1D).

This change also inverts the correlation between BCG immunization and tuberculosis incidence (Table 1). The inclusion of additional global variables highlighted aspects already observed in medical clinics, such as positive correlations between the percentage of the population with a BMI greater than 25 and COVID-19 death and case rates (Table 1).

**Table 1.** Correlations between other variables of interest.

| Dataset | Correlation variables | Correlation method | Correlation value | p-value |
| --- | --- | --- | --- | --- |
|  | COVID-19 case rate and incidence of BMI > 25 | Spearman | 0.57 | 1.4E-16 |
|  |  | Pearson | 0.39 | 9.5E-8 |
|  | COVID-19 death rate and incidence of BMI > 25 | Spearman | 0.56 | 6.0E-16 |
|  |  | Pearson | 0.28 | 1.8E-4 |
|  | COVID-19 case rate and life expectancy in 2019 | Spearman | 0.70 | 4.1E-28 |
|  |  | Pearson | 0.48 | 8.3E-12 |
|  | COVID-19 death rate and life expectancy in 2019 | Spearman | 0.67 | 9.2E-25 |
|  |  | Pearson | 0.38 | 1.5E-7 |
| 1yo BCG immunization rates without replacement | Immunization rate in 2000 vs Estimate incidence of Tuberculosis (per 100k people) in 2018 | Spearman | -0.32 | 5.2E-5 |
|  |  | Pearson | -0.10 | 0.20 |
| 1yo BCG immunization rates introducing 0% for NA data | Immunization rate in 2000 vs Estimate incidence of Tuberculosis (per 100k people) in 2018 | Spearman | 0.19 | 7.7E-3 |
|  |  | Pearson | 0.22 | 2.2E-3 |
| General coverage without replacement | Immunization rate in 2000 vs Estimate incidence of Tuberculosis (per 100k people) in 2018 | Spearman | -0.41 | 4.4E-7 |
|  |  | Pearson | -0.38 | 3.2E-6 |
| General coverage introducing 0% for NA data | Immunization rate in 2000 vs Estimate incidence of Tuberculosis (per 100k people) in 2018 | Spearman | 0.19 | 7.7E-3 |
|  |  | Pearson | 0.12 | 9.2E-2 |

Our analyses did not include any confounding factors that affect the evolution of the outbreak (e.g. implementation of social distancing measures, demographics or other social determinants of health).

Our results expose the limitations of using different analytical approaches and partial datasets in the context of an ongoing outbreak, illustrating how data can be interpreted very differently. Apart from any biological conclusions from our analytical approach regarding the BCG immunization rate, these limitations should be considered before making definitive associations based on correlations that may change over the time of an outbreak.

## Data Availability

Data used in this analyses were obtained from public data repositories from the links below on 17 th may 2020.

https://www.who.int/data/maternal-newborn-child-adolescent/indicator-explorer-new/mca/baccille-calmette-gu%C3%A9rin-(bcg)-immunization-coverage-among-1-year-olds-(-)

https://www.ecdc.europa.eu/en/publications-data/download-todays-data-geographic-distribution-covid-19-cases-worldwide

https://www.who.int/immunization/monitoring_surveillance/data/en/

## Acknowledgements

This research was supported in part by the Biotechnology and Biological Sciences Research Council (grant BB/S006281 to MAV & GLC)

